# From Adversity To Psychopathology: Long-Term Epigenetic Consequences in Adversity-Divergent Twins

**DOI:** 10.1101/2025.03.28.25324831

**Authors:** Dominika Repcikova, Jeanne Le Cléac’h, Archibold Mposhi, Jonathan D. Turner, Conchita D’Ambrosio, Claus Vögele

## Abstract

This study aims to improve our understanding of how previous life experiences (psychosocial adversity, PSA) affect mental health through biological mechanisms. We examine how PSA affects DNA methylation (DNAm) patterns, and its implications for mental health. Using a sample of monozygotic (MZ) twins, this study controls for confounding genetic factors, focusing exclusively on PSA experienced postnatally. Data on PSA is collected using questionnaires, while psychological symptoms are assessed with a structured clinical interview. DNAm data is derived from whole blood. We will first assess whether variations in DNAm mediate the link between PSA and within-pair differences in psychological symptoms and their severity. We will then evaluate hypo- and hypermethylation in selected genomic regions and explore potential associations with psychological symptoms. Finally, we hypothesize that genes exhibiting differential methylation in regulatory regions influence mental health outcomes.

## Introduction

Mental ill-health is one of the leading global challenges, affecting millions of individuals across all socioeconomic backgrounds. Depression, anxiety, and other mental disorders contribute significantly to the global burden of disease, with mental disorders being among the primary causes of disability worldwide^1^. These disorders are characterized by disruptions in emotion regulation, cognitive function, and social behaviors, which can profoundly affect an individual’s quality of life^2^. This highlights the need for a better understanding of the etiology of mental disorders, to target high-risk groups and establish preventative measures. The link between life adversity and mental ill-health is well-documented in the literature; however, the identification and understanding of the mediating factors and mechanisms remain an ongoing area of research.

Adversity can be conceptualized as a series of life events, that pose significant challenges or threats to an individual’s wellbeing. These can include physical or emotional abuse, neglect, peer rejection, household dysfunction and conflict, and broader socio-economic disadvantages, experienced particularly in childhood^3^. Similarly, according to a broader definition, stress occurs when a threat, that is perceived as uncontrollable or unpredictable, is critical to an organism’s wellbeing^4^. There is compelling data showingthat exposure to adversity and stress in childhood and early adulthood affect mental health outcomes, but not all underlying mechanisms are well understood.

In an effort to identify some of the factors contributing to the interplay between adversity and the etiology of mental disorders, epigenetics has emerged as a pivotal field. Epigenetic modifications such as DNA methylation, histone modification, and non-coding RNA interactions, are dynamic processes influenced by environmental factors and life experiences^5^. Adverse life events in childhood such as abuse or trauma can modify the epigenome via DNA methylation^6^. This makes DNA methylation a potential mediating factor between adversity and mental disorders. Epigenetic modifications, including DNA methylation, are associated with inflammaging and immunesenescence, promoting a chronic, low-grade inflammatory systematic state^7^. These immune changes can disrupt brain function through neuroinflammation^8^, HPA axis dysregulation^9^, and neurotransmitter imbalances^10^, thereby contributing to the development and progression of mental disorders. Specifically, DNA methylation has been implicated as a biomarker in a broad spectrum of mental disorders including major depression, bipolar disorder, schizophrenia, and alcohol dependence disorder^6^. DNA methylation, specifically occurring at cytosine-phosphate-guanine (CpG) sites in regulatory regions plays a critical role in modulating gene expression^11^. CpG methylation can silence gene expression by hindering the binding of transcription factors to DNA or shaping a DNA structure favourable to the expression of other genes^11^. This can lead to dysregulation of genes essential for neuronal function, potentially explaining the link with mental disorders^11^.These mechanisms provide a model for understanding how adverse life events contribute to the onset and severity of psychological symptoms.

DNA methylation patterns are particularly responsive to external factors from conception throughout prenatal development. Multiple maternal lifestyle factors can play a role such as stress, smoking, or taking various nutritional supplements making it challenging to compare adversity-induced methylation patterns across individuals^6^. To isolate methylation effects that happened exclusively in childhood, adolescence, and early adulthood we took blood samples from 9 pairs of monozygotic (MZ) twins (N=18), and in addition 4 unpaired blood samples. To investigate the role of adversity-induced DNA methylation in the development of psychological symptoms, we selected a sample of twin pairs with unequal experiences of adversity across various stages of their life. The aim of this study is to examine which CpG methylation sites or patterns and their associated genes act as mediating factors in the interplay of PSA and the development of mental disorders.

Additionally, we are interested in understanding how the level of DNA methylation is reflected in the severity of psychological symptoms, contributing to a scale model of mental disorder. To the best of our knowledge, no study has comprehensively examined the broad range of mental disorders outlined in the Diagnostic and Statistical Manual of Mental Disorders, Fifth Edition (DSM-5)^12^, in relation to adversity-induced DNA methylation as a mediating factor, while accounting for other contributing factors beyond adversity. Building on prior findings, we hypothesize that the link between differences in the experience of adversity, and differences in psychological outcomes in each twin pair will be mediated by variations in DNA methylation patterns (Hypothesis 1). We further hypothesize that DNA hypo- or hypermethylation will be associated with increased or decreased psychological symptoms that can be mediated by changes in gene regulation (Hypothesis 2). Finally, we hypothesize that differentially methylated genes are implicated in psychological outcomes (Hypothesis 3). Our null hypotheses pose that there is no mediation effect of adversity-induced CpG methylation on psychological symptoms, and that the level of CpG methylation is not reflected in the severity of reported psychological symptoms, and that no effect of methylation can be observed in genes implicated in mental health. Our results have the potential to inform at-risk populations about interventions to counter the effects of immuneaging and immunosenescence that emerge as a result of PSA, leading to an overall improvement in their mental health outcomes.

## Methods

### Ethics information

This study was approved by the respective ethics committees of the University of Bielefeld (approval number: EUB2020-184) and the University of Luxembourg (approval number: ERP 22-078). All participants provided informed consent and gave permission for data processing and retention under GDPR prior to the study. Participants were also informed that they can withdraw from the study at any time without stating a reason and request the deletion of their data as long as it has not been included in the aggregate statistics. All collected data underwent pseudonymization, and no individual will be identifiable in any published results. All participants received a compensation of 40€ for providing their biological samples, and 50€ for participating in the clinical psychological interviews.

### Pilot data

We hypothesize that the experience of adverse life events, as defined in the literature^13–17^, including serious health events, job loss, reductions in income, social rejection, bullying, financial struggles, and difficulties with family or housing situations, differs significantly between MZ and DZ twin pairs. Evidence from prior studies demonstrates that even within twin pairs residing in the same household, these experiences differ in terms of perception. Such differences in the experience and salience of adverse life events emerge during early adolescence and increase into early adulthood^18^. Moreover, given that twin-specific differences in socioeconomic status (SES) persist throughout the lifespan, these differences are reflected in the divergence of twins’ epigenomic profiles with age^19–21^.

The relationship between psychosocial stress and epigenetic changes, particularly through DNA methylation patterns, has been well established in the literature^22^. Exploiting the genetic similarity of MZ twins allows us to isolate environmentally induced pathophysiological differences within twin pairs. By focusing on psychosocial stress as a key environmental factor, we aim to identify exposures that may drive epigenetic modifications.

### Design

All participants will be twin pairs sampled from the 3 oldest waves of the TwinLife study (N=710, mnage_cohort2_ = 18.0, mnage_cohort3_ = 24.1, mnage_cohort4_ = 30.0) REF, belonging to a genetic study investigating the socioeconomic and health inequalities across multiple stages of life^23,24^.To investigate the hypotheses outlined in this study, in phase 1 we will assess socioeconomic indicators of childhood, adolescence, and early adulthood adversity, using selected items from the Humiliation Scale^25^, Other as Shamer Scale – 2^26^, Brief Daily Stressors Screening Tool^27^, and Social Support Scale^28^.These questionnaires explore perceived social rejection, financial difficulty, familial and household conflict, and job dissatisfaction. In addition, we will screen participants on the occurrence of events that are generally perceived as stressors, such as illness/injury in the family, own illness/injury, death in the family, job loss, or separation/divorce. Participants will be asked to indicate whether the stressors outlined in the questionnaires took place and if yes, at which life stage, and to rate the perceived negative impact from these stressors on a 5-point Likert scale. Twin pairs who completed the full pre-screening and whose within-pair total score difference was at least 1 standard deviation, will be contacted for subsequent biological sampling and clinical psychological assessment. Participants who expressed interest in phases 2 (biological sampling) and 3 (clinical psychological assessment) of the study will receive instructions for blood and saliva collection at their respective general practitioners’ offices. For the purposes of the study, we will only collect biological data from MZ twins, as we are primarily interested in their DNAm level within-pair differences. DNA extracted from whole blood collected in Paxgene blood RNA tubes will be used to assess DNAm using the Infinium Methylation EPIC v2.0 BeadChip (Illumina). Mental disorders will be assessed using an adjusted version of the Mini-DIPS interview in German language^29^, screening participants for symptoms related to anxiety disorders, affective disorders, eating disorders, sleep-wake disorders, somatic symptom disorders, substance abuse disorders, and obsessive-compulsive disorders.

Our preliminary results show strong positive correlations between between life adversity scores and psychological symptom scores, in both population level and within-pair deltas. Additionally, we will investigate if this is reflected in the changes in DNAm.

#### Phase 1

To establish the degree of perceived social hardships and adversity growing up, we propose to collect the results obtained from our pre-screening questionnaire and summarize them as standardized scores. Subsequently we will calculate within-pair deltas using a tripartite approach of absolute differences between standardized scores, Euclidean distances, and standard deviations within each pair as sanity check.

#### Phase 2

Participants will be asked to set the biosampling on a Tuesday or a Wednesday and communicate the location, date and time of the appointment to facilitate transport and sample processing afterwards. Participants will be asked not to eat 30min prior biosampling and postpone the appointment in case of acute inflammation/infection to not bias data. Blood of the selected MZ Immunotwin participants will be collected at local medical facilities or at home by certified doctors or nurses employing sterile techniques to draw circulatory blood. Peripheral blood will be collected into Paxgene Blood RNA Tubes (BD Biosciences, Franklin Lakes), preserving RNA integrity during transportation and storage, and 10ml into EDTA blood tube (KS Medical, Seoul). Sealed tubes will be kept at 4°C prior to being securely packaged and transported at 4°C to the Luxembourg Institute of Health (LIH) within the next 19h to 57h post-biosampling. Upon arrival at LIH, 2mL of blood of the PaxGene RNA tubes will be aliquoted in two 1mL tubes and stored in accordance with supplier recommendations 24h at −20°C and −80°C for long-term storage. The EDTA tubes will be kept at 4°C between experiments and processed on the day of arrival for Peripheral Blood Mononuclear Cells (PBMC) and plasma isolation as well as flow and mass cytometry.

Data collection was performed blind to the conditions of the experiments. DNA isolation will be performed on 22 blood samples, out of which 18 belong to 9 twin pairs, and 4 samples are unpaired, stored in PaxGene RNA tubes (BD Biosciences, Franklin Lakes). The DNA extraction protocol is based on QIAamp DNA Blood Midi Kit (Qiagen, Hilden) recommendations although some adjustments were made. 2ml of PaxGene (BD Biosciences) whole blood mix per sample will be used. The samples will be centrifuged at 3894g for 10min after thawing at room temperature (RT). The pellet will be washed, in 1ml water for centrifugation at 3894g for 10min, and resuspended in 1ml of PBS (Phosphate Buffered Saline) before starting the procedure. For this experiment, Qiagen protease will be replaced by 25ul proteinase K (Qiagen) per sample incubating for 10min at 56°C. DNA quality will be assessed by Nanodrop (ThermoFisher, Eugene), quantified with Qubit dsDNA HS Assay Kit (ThermoFisher) and stored at −20°C until bisulfite conversion. 350 to 500ng of DNA will be used for bisulfite conversion following the EZ DNA Methylation Kit (Zymo research, Irvine, California, US) recommandations.

To assess DNA methylation patterns we will use the Infinium Methylation EPIC v2.0 BeadChip (Illumina) and iScan (Illumina). Samples will be processed according to the Infinium HD Methylation Assay Reference Guide’s recommendations (Illumina) using GenomeStudio Software 2.0 (Illumina) for quality control. Participants’ data will be processed in 3 experimental batches.

Data will be analysed with R using SeSAMe pipeline (Sensible step-wise analysis of DNA methylation bead chips)^30^ for pre-processing as recommended by Illumina. Data filtering will be done to remove probes that failed the reach the minimum threshold based on our selection criteria. β –values ranging from 0 to 1 will be used for further analysis where a value of 0 indicates that none of the copies of a specific CpG site are methylated, while a value of 1 means that all copies are methylated^31^.

For within-pair differential analysis, delta of β-values will be calculated, reflecting the differential methylation in each twin pairs for all CpG sites.

#### Phase 3

Clinical-psychological interviews conducted online will provide psychological symptom scores. We will blind this phase in a way that the interviewer is not aware of any of the participant’s other scores and CpG methylation levels prior to or during the interviews. Each participant follows the same item protocol based on the German version of the Mini-DIPS clinical interview^29^, in which we map the presence, severity, duration, and frequency of each psychological symptom as defined in the DSM-5^12^. Participants attend the clinical interviews in random order. Items can be either answered on a 1-0 scale (symptom present or absent) or as a number indicating the severity, duration, or frequency of each symptom. In some instances, participants may report unrealistically high frequencies with which they experienced symptoms (e.g. 100 times per month). To statistically control for these outliers, we will cap scores at each symptom group’s respective scale maximum. Symptom scores will be then calculated across all symptom groups for every individual participant, and deltas will be calculated as absolute differences between every sibling in a pair. We will also summarize items that are indicative of the presence of a disorder at a clinical level, as defined by the DSM-5, without accounting for symptom severity.

As these deltas are unstandardized scores without a defined cutoff, we opt for linear regression analysis rather than comparing means between experimental groups.

## Data Availability

All data produced in the present study are available upon reasonable request to the authors

---

**Table 1.** Design Table.

| Question | Hypothesis | Sampling plan<br>(e.g. power analysis) | Analysis Plan | Interpretation given to different outcomes |
| --- | --- | --- | --- | --- |
| Does CpG DNA methylation act as a mediating factor between adversity and mental ill-health? | DNAm mediates the association between variations in psychosocial adversity, and variations in psychological symptoms in MZ twin pairs. |  | High-dimensional mediation analysis (HIMA) and Lavaan analysis on the mediation effect and its size of CpG methylation level, adversity screening score, and psychological symptom score. | Other outcome than H1 might be due to small sample size or incorrect assessment of adversity or psychological symptoms, or due to the fact that there is no mediation effect of CpG methylation on the relationship between adversity and psychological symptoms. |
| Is the level of DNA methylation reflected in psychological symptoms? | DNA hypo- or hyper-methylation will be associated with increased or decreased psychological symptoms that can be mediated by changes in gene regulation. |  | Linear regression between within-pair delta in CpG methylation level and within-pair delta in psychological symptom score and CpG sites association with genes and possible biological pathways (geneOntology analysis). | Other outcome than H2 might be due to small sample size or incorrect assessment of adversity or psychological symptoms or absence of mediation effect of methylation level on psychological symptoms intensity. |
| Which genes affected by hypo- or hypermethylation are implicated in psychological symptoms? | Genes differentially methylated in regulatory regions will have an impact on outcomes in psychological health. |  | Bivariate correlation, ANOVA, and Wilcoxon paired-samples test between siblings, and on variables: DNA methylation, and psychological symptom severity | Other outcome than H3 might be due to small sample size. |

## Supplementary information

Pilot data is reported in

Turner, J. D., D’Ambrosio, C., Vögele, C. & Diewald, M. Twin Research in the Post-Genomic Era: Dissecting the Pathophysiological Effects of Adversity and the Social Environment. *Int. J. Mol. Sci*. **21**, 3142 (2020).

## Acknowledgements

J.D.T. and C.V. were funded by the Fonds National de Recherche (FNR) Luxembourg (C12/BM/3985792 “EpiPath”); J.D.T., C.V., and C.D. are funded by the FNR (C19/SC/13650569), J.D.T. was furthermore funded by the FNR (C16/BM/11342695 “MetCOEPs”; INTER/ANR/16/11568350 “MADAM”). J.D.T. is a management board member of the EU-funded COST actions CA18211 and CA16120.

The funders have/had no role in study design, data collection and analysis, decision to publish or preparation of the manuscript.

## Author contributions

Conceptualisation. J.D.T., C.D., C.V.; Methodology, J.D.T., C.D., C.V., D.R., J.C., A.M.; Resources and formal analysis, J.D.T., C.V.; Writing—original draft, D.R., J.C., A.M.; Writing—Review & Editing, D.R., J.C., A.M., J.D.T., C.V. All authors have read and agreed to the published version of the manuscript.

## Competing interests

The authors declare no competing interests.

## Notes

### Competing Interest Statement

The authors have declared no competing interest.

### Author Declarations

This study was approved by the respective ethics committees of the University of Bielefeld (approval number: EUB2020-184) and the University of Luxembourg (approval number: ERP 22-078). All participants provided informed consent and gave permission for data processing and retention under GDPR prior to the study. Participants were also informed that they can withdraw from the study at any time without stating a reason and request the deletion of their data as long as it has not been included in the aggregate statistics. All collected data underwent pseudonymization, and no individual will be identifiable in any published results. All participants received a compensation of EUR40 for providing their biological samples, and EUR50 for participating in the clinical psychological interviews.

